# Evaluating the impact of Advice and Guidance on diagnostic pathways and healthcare use in England: retrospective cohort study, 2015-2022

**DOI:** 10.64898/2026.10.08.26365047

**Authors:** Kayleigh J Mason, Victoria K Welsh, Kelvin P Jordan, James Bailey, Ram Bajpai, Lorna E Clarson, Alice Faux-Nightingale, Tina Hadley-Barrows, John K Haines, Rosie Harrison, Toby Helliwell, Samantha L Hider, Clare Jinks, Natalie Knight, Christian D Mallen, Claire Burton

## Abstract

**Objective:** To determine outcomes by referral route (any Advice and Guidance [A&G] versus elective referrals only) for patients with inflammatory bowel disease (IBD), Parkinson’s disease, and systemic lupus erythematosus (SLE).

**Design:** Population-based retrospective cohort study.

**Setting:** Linked Clinical Practice Research Datalink (CPRD) Aurum primary care records, 2015-2022.

**Participants:** Patients with incident diagnosis of IBD, Parkinson’s disease, or SLE.

**Main Outcome Measures:** Time from the earliest recorded A&G or elective referral to diagnosis, first recorded definitive treatment and first attended outpatient appointment. Secondary outcomes included counts of primary care consultations, prescriptions, referrals, and outpatient appointments. Propensity score weighted flexible parametric survival analyses accounted for differences between exposure groups.

**Results:** Among 71,145 eligible patients, A&G was recorded for 1,137 (3.0%) with IBD, 971 (3.4%) with Parkinson’s, and 243 (4.9%) patients with SLE. Compared with elective referral only, A&G was associated with slower time to diagnosis for IBD (adjusted median difference 15.9 weeks; hazard ratio 0.58, 95% confidence interval 0.52 to 0.66), Parkinson’s disease (11.4 weeks; 0.57, 0.51 to 0.64), and SLE (6.3 weeks; 0.82, 0.73 to 0.91). However, A&G was not associated with a longer time to treatment or first attended outpatient appointment. An increase in primary care consultations following A&G was observed in patients later diagnosed with IBD and SLE.

**Conclusion:** A&G was associated with a longer diagnostic interval than elective referral only. These findings do not establish that A&G caused diagnostic delay. A&G might be used at an earlier stage of more complex diagnostic pathways.

**What is already known on this topic:**

- Advice and Guidance (A&G) enables primary care clinicians to obtain specialist input before or instead of elective referral and is promoted to support patient management and reduce unnecessary outpatient activity.
- Evidence is limited on whether A&G shortens diagnostic and treatment pathways or instead adds an additional step for patients who ultimately require specialist care.
- The effects of A&G on subsequent primary care workload, secondary care use, and patient safety are uncertain, and implementation varies between local healthcare systems.

**What this study adds:**

- Among patients subsequently diagnosed with inflammatory bowel disease (IBD), Parkison’s disease or systemic lupus erythematosus (SLE), the interval from the earliest recorded A&G or elective referral of ***any type*** to diagnosis was longer among those with any recorded A&G although we did not observe an increase time to treatment or first attended outpatient appointment.
- Patients with A&G had more ***all-cause*** subsequent referrals across all three conditions and more ***all-cause*** primary care consultations for IBD and SLE; no clear differences were found in attended outpatient appointments, unplanned hospital admission, or mortality.
- A&G may identify an earlier stage of the pathway; however, patients recorded with A&G tended to be more complex and findings should be interpreted as associations rather than evidence that A&G caused delay.

## Introduction

Referral from primary to secondary care is a central function of UK general practice and a key route through which patients access specialist assessment, diagnosis, and treatment.[1–3] In recent years, Advice and Guidance (A&G) has become increasingly prominent and enables primary care clinicians to seek specialist input through a formal recorded electronic process, with the intention of supporting management in the most appropriate setting, reducing unnecessary outpatient activity, and improving the efficiency of referral pathways.[4,5]

The COVID-19 pandemic accelerated changes at the interface between primary and secondary care, including increased use of remote, electronic, and triage-based approaches to specialist input;[6,7] A&G has subsequently been promoted as a mechanism to support elective recovery and manage waiting list pressures.[8,9] However, while A&G may shorten some patient journeys and enable earlier specialist input,[10–12] concerns remain about whether it could also delay diagnosis or treatment for patients who ultimately require specialist assessment.[13,14] Evidence on the effectiveness, safety, and equity of A&G remains limited, particularly when considered from the perspective of patient outcomes and subsequent healthcare utilisation.[15] To this end a recent report by the Health Services Safety Investigations Board (HSSIB) concluded that A&G can improve access to specialist expertise and support care closer to home, but its implementation has been inconsistent and, in some cases, has contributed to patient harm.[15]

Existing studies suggest that A&G can be efficient in selected clinical settings, but the impact of A&G is likely to depend on case mix, specialty, local implementation, and the extent to which advice substitutes for, delays, or accelerates specialist review.[5,10,11] Important uncertainties remain around which patients are suitable for A&G, how frequently A&G results in subsequent referral, whether access differs by sociodemographic or clinical characteristics, and whether use of A&G affects downstream demand in primary and secondary care.[4,15]

Inflammatory bowel disease (IBD), Parkinson’s disease, and systemic lupus erythematosus (SLE) were selected through primary care clinician interviews [5], stakeholder and patient consensus as clinically distinct tracer conditions requiring specialist input, with measurable diagnostic and treatment pathways in routine data.

The primary objective was to compare time to diagnosis, time to definitive treatment and time to first outpatient appointment among patients with a recorded A&G request before diagnosis and those with elective referral only. The secondary objective was to compare subsequent healthcare utilisation, including primary care consultations, prescriptions, referrals, outpatient appointments, unplanned hospital admissions, and mortality.

## Methods

### Study design and data sources

We undertook a retrospective cohort study using CPRD Aurum primary care data. CPRD Aurum is a UK representative database of anonymised primary care electronic health records for over 50 million patients from over 1,800 general practices in England using EMIS Web® software, covering 25% of the UK population.[16] CPRD Aurum was linked at patient level, where available, to Hospital Episode Statistics (HES) Admitted Patient Care, HES Outpatients, Office for National Statistics (ONS) death registration data, CPRD ethnicity records, patient-level Index of Multiple Deprivation, and practice-level Clinical Commissioning Group pseudonyms (referred to as commissioning localities).

The study received approval from CPRD Research Data Governance (#25_005032) and used the June 2025 release.[16]

### Study population

The study population comprised all eligible individuals in CPRD Aurum with a first recorded diagnosis of one of three tracer conditions: inflammatory bowel disease (IBD), Parkinson’s disease, or systemic lupus erythematosus (SLE). These conditions were selected through a multisource consensus process involving interviews with primary care clinicians (asked about conditions/specialties where A&G was used to expediate diagnosis/treatment; [5]), stakeholders, and patient and public involvement and engagement (PPIE) contributors. They were selected because IBD provides broad age coverage of the gastroenterology referral pathway, Parkinson’s disease provides an older adult neurological referral pathway, and SLE is a clinically important, multi-system condition spanning several specialties with a more diverse, middle-aged female population. Further details on the consensus process and selection criteria are provided in Supplementary Table S1.

Code lists for tracer conditions, A&G and elective referrals, and covariates are available here: https://doi.org/10.21252/8rgt-5w70

The study period included diagnoses recorded between 1 January 2015 and 31 December 2022. Individuals were required to have at least 24 months of registration history in CPRD Aurum before the first recorded diagnosis date.

The index diagnosis date was defined as the earliest recorded diagnosis date across CPRD Aurum, Hospital Episode Statistics Outpatients, or Hospital Episode Statistics Admitted Patient Care. Analyses were stratified by tracer condition.

### Exposure definition

The exposure of interest was the recorded referral pathway in the 24 months (aligned with registration history) before the index diagnosis date. Patients were classified into the following groups:

- **Any A&G group:** patients with any recorded A&G event in the 24 months before diagnosis. Patients with both A&G and elective referral recorded during this period were classified in the A&G group.
- **Elective referral only group:** patients with elective referral recorded in the 24 months before diagnosis and no recorded A&G event.
- **No recorded A&G or elective referral group:** patients with neither A&G nor elective referral recorded in the 24 months before diagnosis. This group was described and included in analyses of subsequent healthcare utilisation where appropriate.

A&G events were determined from coded primary care records using terms including “*Advice and guidance requested*” and “*Advice and guidance received*”. Elective referrals were determined using coded referral terms indicating routine referral to a specialty, excluding fast-track referrals, emergency referrals, and referrals with an intention to admit.

### Outcomes

The primary time-to-event outcomes are:

- time from the earliest recorded A&G or elective referral event to diagnosis of the tracer condition,
- time from earliest recorded A&G or elective referral to first recorded definitive medical treatment for the tracer condition, and
- time from earliest recorded A&G or elective referral to first hospital outpatient appointment.

The earliest recorded event could be an elective referral in the any A&G group as they were classified based on ever being recorded with A&G.

Definitive medical treatment was defined using condition-specific therapy lists (Supplementary Table S2). For IBD, treatments included anti-inflammatories, corticosteroids, immunomodulators, biologics, and Janus kinase inhibitors. Treatments for Parkinson’s disease included dopamine precursors, dopamine agonists, and monoamine oxidase-B inhibitors with treatments for SLE including corticosteroids, immunomodulators, biologics, and human immunoglobulin.

Secondary healthcare utilisation outcomes were measured during the 24 months after the earliest recorded A&G or elective referral event and included:

- number of primary care consultations (defined as the number of different days with a healthcare practitioner contact),
- number of unique drug substances prescribed in primary care,
- number of referrals recorded in primary care,
- number of attended outpatient appointments recorded in Hospital Episode Statistics Outpatients,
- an all-cause overnight unplanned hospital admission recorded in Hospital Episode Statistics Admitted Patient Care, and
- time to all-cause mortality using Office for National Statistics death registration data.

### Covariates

Covariates were measured on or within 24 months before the earliest recorded A&G or elective referral event including age bands (0-14, 15-24, 25-34, 35-44, 45-54, 55-64, 65-74, 75-84 and 85+ years), gender, geographical region, ethnicity (using CPRD Aurum Ethnicity algorithm; [17]), deprivation (patient-level Index of Multiple Deprivation quintile; [18]), multimorbidity (Cambridge Multimorbidity Score; [19,20]), polypharmacy (number of unique drug substances prescribed), year of referral, and time to diagnosis (only for models of post-referral healthcare utilisation).

### Statistical analysis

Baseline characteristics were summarised by referral pathway using frequencies and percentages for categorical variables, and medians with 25th-75th percentiles for continuous variables. Small cell counts (≤5) were suppressed in outputs to protect confidentiality. Outputs were checked to ensure suppressed counts cannot be derived by subtraction or other manipulation.

Absence of a code for A&G or elective referral was assumed to indicate no such an event had occurred. All included individuals had year of birth (to calculate age), consultation year and gender recorded. Individuals without data for ethnicity, deprivation or commissioning locality were recorded as “unknown”.

Analyses were conducted separately for each tracer condition.

For the primary outcomes, we estimated the median time in weeks (25th-75th percentiles) for those recorded with the outcomes. Flexible parametric survival models compared time to diagnosis between patients with any recorded A&G and those with elective referral only. Flexible parametric models were selected because they allow flexible modelling of the baseline hazard, accommodate time-dependent effects, and facilitate estimation of absolute as well as relative effects [21]. Standardised conditional medians were used to compare estimated population-average survival times in weeks for individuals recorded with any A&G or elective referral only, while retaining the observed distribution of propensity scores.

Propensity score weighting balanced the A&G and elective referral only groups on measured covariates. The propensity score was estimated using logistic regression and trimmed between 0.01-0.99 to omit extreme weights and reduce bias from unmeasured confounders.[22] Candidate covariates included gender, age bands, deprivation, ethnicity, Cambridge Multimorbidity Score, polypharmacy, commissioning locality, and calendar year of tracer condition diagnosis. Balance after weighting was assessed using estimated bias across deciles of the propensity score (Supplementary Figure S1). Unadjusted and propensity score-weighted hazard ratios with 95% confidence intervals are reported.

The same approach was used for the secondary time-to-event outcome of death.

For healthcare utilisation outcomes measured in the 24 months after the earliest A&G or elective referral event, negative binomial regression was used to estimate incidence rate ratios for counts of primary care consultations, prescriptions, referrals, and outpatient appointments. Robust Poisson regression was used to compare the two group on the proportion with at least one all-cause unplanned hospital admission.

Following the adjusted model, marginal predictions were used to compare estimated population-average expected counts (for primary care consultations, prescriptions, referrals, and outpatient appointments) or proportion with unplanned hospital admissions within each referral pathway, standardised to the observed distribution of propensity-score categories.

Robust standard errors were used to account for clustering within general practices.

Data management and statistical analyses were performed using Stata MP version 19.5 (StataCorp, USA).

This study meets all five of the CODE-EHR minimum framework standards for the use of structured healthcare data in clinical research, with one preferred criterion also met (Ethics and Governance). Full details are provided in the completed CODE-EHR checklist (Supplementary Table S3).[23]

### Patient and public involvement

PPIE informed the design of the study, including the selection of tracer conditions and of outcomes related to patient burden and healthcare use. A PPIE co-applicant is a member of the study team and contributed to protocol development, interpretation of findings, and dissemination. A dedicated PPIE group met regularly and advised on methodology, interpretation, and dissemination. Additionally, the study stakeholder group (n=7) advised on clinical relevance (1x GP; 1x consultant), ethics (1x senior lecturer), NHS implementation (1x commissioner), stakeholder engagement (1x Healthwatch; 1x knowledge mobilisation lead), and patient priorities (1x public co-applicant), and provided insight into the tracer conditions and outcomes.

### Ethics and governance

The study used routinely collected anonymised data from CPRD Aurum and linked datasets. The study used routinely collected pseudonymised electronic health records only. No additional information was requested from general practices, and no patients were contacted.

## Results

### Study population

Between 1 January 2015 and 31 December 2022, 71,145 individuals in CPRD Aurum had a first recorded diagnosis of one of the three tracer conditions and met the study eligibility criteria. Of these, 37,823 had IBD, 28,341 had Parkinson’s disease, and 4,981 had SLE (Figure 1).

In the 24 months before diagnosis, the distribution of referral pathways varied by tracer condition with A&G recorded in 1,137 (3.0%) with IBD, 971 (3.4%) patients with Parkinson’s disease, and 243 (4.9%) with SLE (Figure 1).

### Baseline characteristics

Baseline characteristics were broadly similar by gender within each tracer condition (IBD females 55% versus 51%, Parkinson’s females 36% versus 38%, and SLE females 83% versus 83% in the any A&G and elective referral only groups, respectively; Table 1).

**Table 1.**
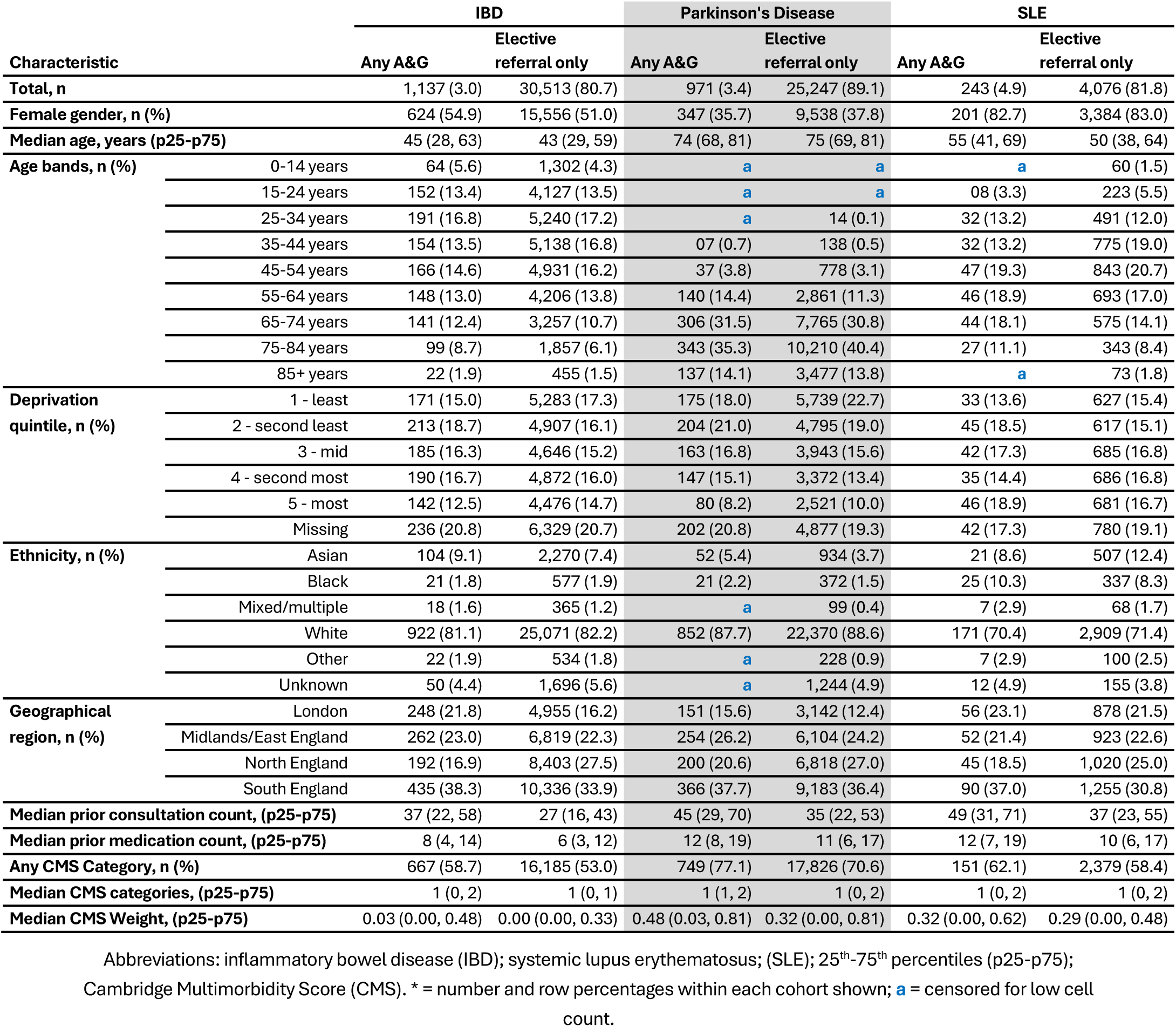
Patient characteristics for those recorded with any A&G or referral only by tracer condition.

Compared with patients with elective referral only, patients with A&G were slightly older for IBD (median 45 vs 43 years) and SLE (55 v 50 years), but marginally younger for Parkinson’s disease (74 vs 75 years).

Deprivation and ethnicity distributions were broadly similar within conditions, although a sizeable proportion of data was missing for deprivation due to a lag in linkage (range 17% v 21%; Table 1). Regionally, patients with A&G were more commonly recorded in South England and London, and less commonly in North England, than patients with elective referral only across the three tracer conditions (Table 1).

Compared with patients with elective referral only, patients with A&G had higher consultation counts in the previous 24 months across all tracer conditions (median 37 vs 27 for IBD, 45 vs 35 for Parkinson’s, and 49 vs 37 for SLE) and slightly higher prescribed medication counts during the same period (8 vs 6, 12 vs 11, and 12 vs 10, respectively). Any record of a condition and/or prescription in the Cambridge Multimorbidity Score was more common among patients with A&G than among those with elective referral only (59% vs 53%, 77% vs 71%, and 62% v 58%, respectively), and median Cambridge Multimorbidity Score was higher in the A&G group across all three tracer conditions (Table 1).

Among patients with no recorded A&G or elective referral in the 24 months before diagnosis, baseline characteristics differed from those with recorded referral activity (any A&G or elective referral only groups) as they generally had fewer prior consultations and medications (Supplementary Table S4). For IBD and SLE, patients without a recorded referral or A&G were younger and had lower recorded multimorbidity, whereas those with Parkinson’s disease were slightly older with greater multimorbidity (median Supplementary Table S4).

### Time to diagnosis, definitive treatment and outpatient appointment

Where hazard ratios are above 1, this should be interpreted as a higher rate of the outcome (e.g., diagnosis) following the referral event, consistent with shorter time to outcome. Where hazard ratios are below 1, this should be interpreted as a lower rate of outcome following the referral event indicating a longer observed interval to outcome.

Any A&G was associated with a longer adjusted time to diagnosis compared with elective referral only for all three conditions after propensity score weighting; the hazard ratio for time to diagnosis associated with A&G was 0.58 (95% CI 0.52, 0.66) for IBD, 0.57 (95% CI 0.51, 0.64) for Parkinson’s disease, and 0.82 (95% CI 0.73, 0.91) for SLE with diagnosis estimated to take an additional 15.9, 11.4 and 6.3 weeks, respectively, for those with any A&G recorded (Table 2).

**Table 2.**
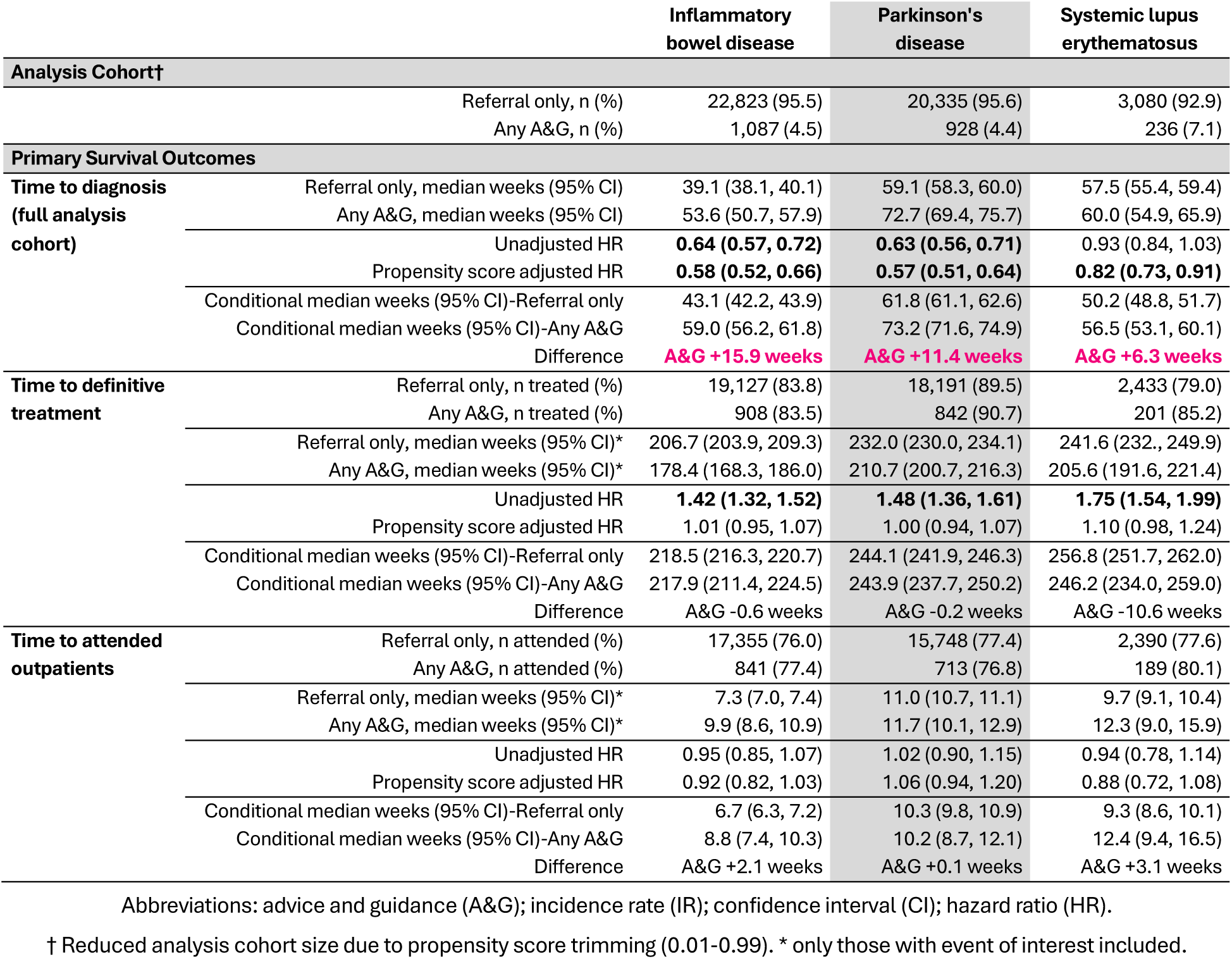
Primary survival outcomes by tracer condition.

Time to first recorded definitive treatment did not differ between referral pathways after propensity score weighting (Table 2). No significant difference in time to first recorded definitive treatment was observed for IBD (1.01; 95% CI 0.95, 1.07), Parkinson’s disease (1.00; 95% CI 0.94, 1.07) or SLE (1.10; 95% CI 0.98, 1.24; Table 2).

Propensity score-weighted analyses showed no clear difference in time to first attended outpatient appointment among patients with A&G compared with elective referral only, with hazard ratios of 0.92 (95% CI 0.82, 1.03), 1.06 (95% CI 0.94, 1.20), and 0.88 (95% CI 0.72, 1.08) for IBD, Parkinson’s disease, and SLE, respectively.

### Healthcare utilisation in the 24 months after referral

In weighted models, A&G was associated with higher rates of primary care consultations for IBD and SLE, primary care prescriptions for IBD, and referrals to secondary care for all three tracer conditions when compared with elective referral only (Table 3).

**Table 3.**
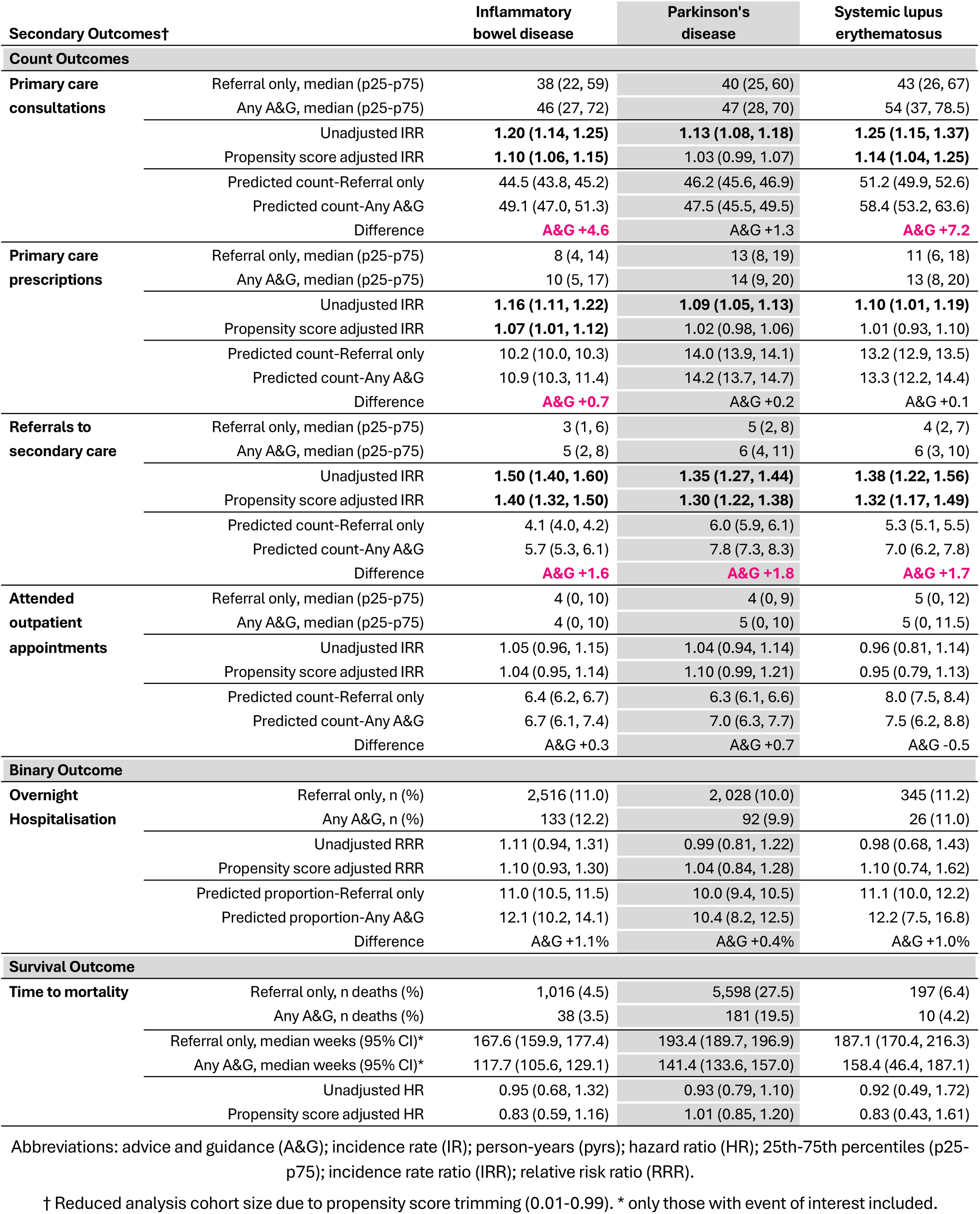
Secondary outcomes by tracer condition.

| Secondary Outcomes† |  | Inflammatory bowel disease | Parkinson's disease | Systemic lupus erythematosus |
| --- | --- | --- | --- | --- |
| **Count Outcomes** |  |  |  |  |
| **Primary care consultations** | Referral only, median (p25-p75) | 38 (22, 59) | 40 (25, 60) | 43 (26, 67) |
|  | Any A&G, median (p25-p75) | 46 (27, 72) | 47 (28, 70) | 54 (37, 78.5) |
|  | Unadjusted IRR | **1.20 (1.14, 1.25)** | **1.13 (1.08, 1.18)** | **1.25 (1.15, 1.37)** |
|  | Propensity score adjusted IRR | **1.10 (1.06, 1.15)** | 1.03 (0.99, 1.07) | **1.14 (1.04, 1.25)** |
|  | Predicted count-Referral only | 44.5 (43.8, 45.2) | 46.2 (45.6, 46.9) | 51.2 (49.9, 52.6) |
|  | Predicted count-Any A&G | 49.1 (47.0, 51.3) | 47.5 (45.5, 49.5) | 58.4 (53.2, 63.6) |
|  | Difference | **A&G +4.6** | A&G +1.3 | **A&G +7.2** |
| **Primary care prescriptions** | Referral only, median (p25-p75) | 8 (4, 14) | 13 (8, 19) | 11 (6, 18) |
|  | Any A&G, median (p25-p75) | 10 (5, 17) | 14 (9, 20) | 13 (8, 20) |
|  | Unadjusted IRR | **1.16 (1.11, 1.22)** | **1.09 (1.05, 1.13)** | **1.10 (1.01, 1.19)** |
|  | Propensity score adjusted IRR | **1.07 (1.01, 1.12)** | 1.02 (0.98, 1.06) | 1.01 (0.93, 1.10) |
|  | Predicted count-Referral only | 10.2 (10.0, 10.3) | 14.0 (13.9, 14.1) | 13.2 (12.9, 13.5) |
|  | Predicted count-Any A&G | 10.9 (10.3, 11.4) | 14.2 (13.7, 14.7) | 13.3 (12.2, 14.4) |
|  | Difference | **A&G +0.7** | A&G +0.2 | A&G +0.1 |
| **Referrals to secondary care** | Referral only, median (p25-p75) | 3 (1, 6) | 5 (2, 8) | 4 (2, 7) |
|  | Any A&G, median (p25-p75) | 5 (2, 8) | 6 (4, 11) | 6 (3, 10) |
|  | Unadjusted IRR | **1.50 (1.40, 1.60)** | **1.35 (1.27, 1.44)** | **1.38 (1.22, 1.56)** |
|  | Propensity score adjusted IRR | **1.40 (1.32, 1.50)** | **1.30 (1.22, 1.38)** | **1.32 (1.17, 1.49)** |
|  | Predicted count-Referral only | 4.1 (4.0, 4.2) | 6.0 (5.9, 6.1) | 5.3 (5.1, 5.5) |
|  | Predicted count-Any A&G | 5.7 (5.3, 6.1) | 7.8 (7.3, 8.3) | 7.0 (6.2, 7.8) |
|  | Difference | **A&G +1.6** | **A&G +1.8** | **A&G +1.7** |
| **Attended outpatient appointments** | Referral only, median (p25-p75) | 4 (0, 10) | 4 (0, 9) | 5 (0, 12) |
|  | Any A&G, median (p25-p75) | 4 (0, 10) | 5 (0, 10) | 5 (0, 11.5) |
|  | Unadjusted IRR | 1.05 (0.96, 1.15) | 1.04 (0.94, 1.14) | 0.96 (0.81, 1.14) |
|  | Propensity score adjusted IRR | 1.04 (0.95, 1.14) | 1.10 (0.99, 1.21) | 0.95 (0.79, 1.13) |
|  | Predicted count-Referral only | 6.4 (6.2, 6.7) | 6.3 (6.1, 6.6) | 8.0 (7.5, 8.4) |
|  | Predicted count-Any A&G | 6.7 (6.1, 7.4) | 7.0 (6.3, 7.7) | 7.5 (6.2, 8.8) |
|  | Difference | A&G +0.3 | A&G +0.7 | A&G -0.5 |
| **Binary Outcome** |  |  |  |  |
| **Overnight Hospitalisation** | Referral only, n (%) | 2,516 (11.0) | 2,028 (10.0) | 345 (11.2) |
|  | Any A&G, n (%) | 133 (12.2) | 92 (9.9) | 26 (11.0) |
|  | Unadjusted RRR | 1.11 (0.94, 1.31) | 0.99 (0.81, 1.22) | 0.98 (0.68, 1.43) |
|  | Propensity score adjusted RRR | 1.10 (0.93, 1.30) | 1.04 (0.84, 1.28) | 1.10 (0.74, 1.62) |
|  | Predicted proportion-Referral only | 11.0 (10.5, 11.5) | 10.0 (9.4, 10.5) | 11.1 (10.0, 12.2) |
|  | Predicted proportion-Any A&G | 12.1 (10.2, 14.1) | 10.4 (8.2, 12.5) | 12.2 (7.5, 16.8) |
|  | Difference | A&G +1.1% | A&G +0.4% | A&G +1.0% |
| **Survival Outcome** |  |  |  |  |
| **Time to mortality** | Referral only, n deaths (%) | 1,016 (4.5) | 5,598 (27.5) | 197 (6.4) |
|  | Any A&G, n deaths (%) | 38 (3.5) | 181 (19.5) | 10 (4.2) |
|  | Referral only, median weeks (95% CI)* | 167.6 (159.9, 177.4) | 193.4 (189.7, 196.9) | 187.1 (170.4, 216.3) |
|  | Any A&G, median weeks (95% CI)* | 117.7 (105.6, 129.1) | 141.4 (133.6, 157.0) | 158.4 (46.4, 187.1) |
|  | Unadjusted HR | 0.95 (0.68, 1.32) | 0.93 (0.79, 1.10) | 0.92 (0.49, 1.72) |
|  | Propensity score adjusted HR | 0.83 (0.59, 1.16) | 1.01 (0.85, 1.20) | 0.83 (0.43, 1.61) |

Abbreviations: advice and guidance (A&G); incidence rate (IR); person-years (pyrs); hazard ratio (HR); 25th-75th percentiles (p25-p75); incidence rate ratio (IRR); relative risk ratio (RRR).
† Reduced analysis cohort size due to propensity score trimming (0.01-0.99). \* only those with event of interest included.

The propensity score weighted incidence rate ratios for primary care consultations were 1.10 (95% CI 1.06, 1.15) for IBD and 1.14 (95% CI 1.04, 1.25) for SLE, with an additional 4.6 and 7.2 consultations predicted for any A&G (Table 3). For prescriptions, weighted incidence rate ratios suggested higher prescribing burden among patients with IBD and A&G compared with elective referral only (propensity weighted IRR 1.07; 95% CI 1.01, 1.12), while similar rates were observed for Parkinson’s disease and SLE (Table 3).

A&G was associated with greater rates of referral to secondary care than those recorded with elective referrals only with propensity score weighted incidence rate ratios of 1.40 (95% CI 1.32, 1.50) for IBD, 1.30 (95% CI 1.22, 1.38) for Parkinson’s disease, and 1.32 (95% CI 1.17, 1.49) for SLE; an additional 1.6, 1.8 and 1.7 referrals were predicted for any A&G, respectively (Table 3).

A&G was associated with similar rates of outpatient appointment attendance (Table 3).

### Unplanned admissions and mortality

Overnight unplanned hospital admissions for any reason occurred in 10%-12% patients across any A&G and elective referral only groups during follow-up. In weighted robust Poisson models, A&G was associated with no clear difference in risk of unplanned admission compared with elective referral only.

Deaths were recorded for 4.4% patients with IBD, 27.2% Parkinson’s and 6.2% SLE. Propensity score weighted flexible parametric survival models showed no clear evidence of difference in all-cause mortality among patients with A&G compared with elective referral only (IBD 0.83, 95% CI 0.59, 1.16; Parkinson’s 1.01, 95% CI 0.85, 1.20; SLE 0.83, 95% CI 0.43, 1.62), although estimates were imprecise for the younger IBD and SLE patient cohorts as there were few events (Table 3).

## Discussion

In this population-based study using linked primary and secondary care records, patients with any recorded A&G in the two years before diagnosis of IBD, Parkinson’s disease and SLE had a longer interval from their earliest recorded referral-related event to diagnosis than patients with elective referral only. This pattern was consistent across the tracer conditions with adjusted differences of approximately sixteen, eleven, and six weeks, respectively. Despite these longer diagnostic intervals, A&G was not associated with a longer time to definitive treatment for any of the three tracer conditions, or in time to first attended outpatient appointment for any condition. A&G was associated with more subsequent referrals to secondary care across all three conditions, and with more primary care consultations for IBD and SLE. Differences in unplanned hospital admission and mortality were not evident.

Taken together, these findings suggest that the use of A&G was not associated with anticipated shortening of diagnostic pathways in patients who received diagnoses of the tracer conditions. A&G may have enabled specialist input without immediately generating an outpatient appointment, but this did not translate into earlier recorded diagnosis or lower subsequent healthcare use.

### Comparison with literature

Evidence on the effects of A&G on complete patient pathways in England remains limited. A systematic review of 72 studies found that electronic consultation was associated with more rapid access to specialist advice and avoidance of some hospital referrals, but most studies were observational, were undertaken in North America, and provided low or very low certainty evidence.[24] Specialty-specific UK reports have also described improvements in communication or outpatient flow, including in neurology and general surgery.[10,11] These studies have evaluated specialist response, referral diversion, or implementation within individual services, rather than time to diagnosis among patients who ultimately require specialist care.

Our findings are consistent with evidence that A&G often operates alongside, rather than instead of, conventional referral. In our previous population-based CPRD Aurum study, 86% of patients with A&G also had an elective referral within four months.[4] The greater number of subsequent referrals observed in the present study might therefore indicate that A&G represented an additional step for some patients, although we could not determine the ordering, purpose, or clinical content of all referral-related events, particularly for A&G.

From patients’ perspectives, diagnostic delay can mean prolonged symptoms, uncertainty, repeated consultations, feeling that concerns are not recognised, and poorer physical and psychological wellbeing. Patients with IBD have described embarrassment, symptoms being mislabelled or not taken seriously, and communication failures across care settings [25]; delays in Parkinson’s disease can prolong the period without an explanation or appropriate treatment [26]; and patients with SLE report frustration, psychological distress, reduced trust in clinicians, and potentially irreversible organ damage [27]. These experiences underline the importance of the longer diagnostic intervals observed with A&G, even if they partly reflect greater clinical complexity rather than an effect of A&G itself.

Differences in case selection could also explain the longer diagnostic intervals. Primary care clinicians report using A&G in response to clinical complexity, uncertainty, difficulties navigating pathways, and a need to validate clinical decisions.[5] The higher previous consultation, prescribing, and multimorbidity burden among patients receiving A&G supports the possibility that these patients had more complex clinical presentations. Although propensity score weighting accounted for measured characteristics, residual confounding by symptoms, disease severity, diagnostic uncertainty, and clinician concern is likely.

The increased primary care activity following A&G might represent both continuing clinical need and implementation of specialist recommendations. Previous electronic consultation research found that primary care clinicians acted on specialist recommendations in most cases, often requiring subsequent communication with patients.[28] Additional consultations or prescriptions should therefore not automatically be considered unnecessary workload. Nevertheless, qualitative evidence from English primary care identifies concern about delayed responses and inequitable transfer of work where specialist recommendations exceed primary care capacity.[5]

The longer observed diagnostic intervals are also relevant to the HSSIB interim evaluation of Advice and Guidance; HSSIB conduct independent investigations of patient safety concerns in the English NHS. HSSIB reported that A&G could improve care when it supported effective communication and timely specialist input.[15] However, incidents involving delayed or missed diagnosis and treatment where processes were poorly implemented or monitored were also identified. There was also limited national data on patient-safety outcomes and variation between local implementation and national expectations.[15] Our observational results cannot determine whether these mechanisms explain the associations found, but they reinforce the need to evaluate whole patient pathways and safety outcomes rather than referral diversion alone.

### Strengths and limitations

This study used a large population-based primary care database linked to hospital admissions, outpatient attendance, deprivation, and mortality data. We examined three tracer conditions with different demographic profiles and specialist pathways, used identical exposure and outcome principles across conditions, and considered diagnostic, treatment, patient-burden, and service-utilisation outcomes between referral pathways. The consistency of the association with time to diagnosis across the three conditions is notable. Patient and public involvement and stakeholders informed the choice of tracer conditions and outcomes.

Our study has several important limitations. The exposure represented the earliest recorded A&G or elective referral during the two years before the tracer condition diagnosis, rather than a confirmed referral for the symptoms that resulted in that diagnosis. A&G records in CPRD Aurum are limited to coded data rather than free text, so do not routinely have the specialty, clinical question, or condition under consideration.[4] Some index events may therefore have related to a different health problem (e.g. were described in the free text under a different or generic problem), particularly among patients with multimorbidity. However, this exposure misclassification could affect both referral groups and the estimated diagnostic intervals and would be unlikely to explain our findings.

Patients with both A&G and elective referral records were classified as having any A&G. The A&G group was therefore heterogeneous and could include patients managed following A&G alone, those referred after A&G, and those who received A&G after an elective referral. Using the earliest event as time zero does not capture the ordering, number, or content of subsequent A&G and referral events. The ordering of A&G and elective referral is likely to be complex, as our previous study showed that elective referral frequently preceded an A&G record.[4] For a patient receiving A&G before referral, the measured interval begins earlier in the diagnostic pathway than it may for a patient whose first recorded event is an elective referral. Consequently, part of the longer interval associated with A&G may reflect the exposure definition and earlier observation of the diagnostic process rather than a delay caused by A&G.

The cohort was defined by a subsequent recorded diagnosis. The findings therefore apply to patients who ultimately received a new diagnosis of IBD, Parkinson’s disease, or SLE, and not to all patients for whom A&G was used. We could not evaluate patients in whom specialist advice appropriately avoided referral or excluded a tracer condition diagnosis. The study consequently cannot estimate the overall effectiveness, efficiency, or safety of A&G as some of its intended benefits may occur among patients who never enter a diagnosed disease cohort.

The referral route was not randomly allocated and therefore the decision to use A&G may have been influenced by symptom severity, diagnostic uncertainty, clinical complexity, patient preference, referral urgency, specialist capacity, local policy, and whether A&G was mandatory.[5] Although propensity score weighting addressed measured demographic, clinical, prescribing, temporal, and locality characteristics, we were unable to account for severity, clinician concern, and reason for using A&G. Residual confounding by indication is therefore likely, and associations should not be interpreted causally.

The absence of a recorded code was assumed to indicate that A&G or elective referral had not occurred. Coding completeness may have varied between practices, clinical systems, localities, and calendar periods. Patients classified as having no recorded A&G or referral had fewer preceding consultations and prescriptions for IBD and SLE, but patients with Parkinson’s disease in this group had greater recorded multimorbidity. This suggests that the group was heterogeneous and may have included patients diagnosed through acute admission, alternative referral routes, private care, or referrals that were not captured in the primary care record.

Primary care prescribing does not fully capture treatment initiated and supplied wholly in secondary care. The date of first prescription in the primary care record could represent transfer of prescribing responsibility rather than treatment initiation. Similarly, outpatient appointments, consultations, prescriptions, and referrals may not have been related to the tracer condition. Mortality analyses had limited precision, particularly for the younger IBD and SLE populations recorded few deaths, and absence of evidence of a difference for mortality and unplanned admissions should not be interpreted as evidence of equivalent safety.

Finally, A&G implementation varied geographically and changed over time, including during the COVID-19 pandemic. Local requirements to use A&G before referral, differences in service capacity, and electronic interoperability could have affected both exposure classification and outcomes. HSSIB has highlighted substantial variation in implementation, pathway oversight, tracking, and management of rejected or redirected referrals[15]. Although weighting included locality and calendar year, it could not capture all relevant variation in local policy or service operation.

### Implications

A&G may improve communication, support primary care management, and prevent unnecessary referral for many patients who do not subsequently receive one of the tracer diagnoses. However, systems should ensure that A&G does not become an additional administrative hurdle for patients requiring specialist assessment, nor an inappropriate work shift from secondary care to primary care.

Further evaluations of A&G should move beyond measures of requests completed or referrals avoided. Core outcomes should include time to definitive diagnosis and treatment, repeated contacts across primary and secondary care, impact of A&G in creating additional workload associated with a patient, conversion to referral, rejected or redirected referrals, unplanned hospital use, patient-reported experience, and safety incidents. These outcomes were determined as important by our patient, public and stakeholder groups. Analyses should distinguish A&G that substitutes for referral from A&G preceding, following, or operating alongside referral. These priorities align with HSSIB’s concerns about variable pathway oversight, interoperability, workforce training, and limited data on patient safety outcomes.[15]

### Conclusion

Among patients subsequently diagnosed with IBD, Parkinson’s disease, or SLE, A&G was consistently associated with a longer interval to diagnosis than elective referral only, however, was not associated with longer time to treatment initiation. A&G was also associated with more subsequent referrals across all three conditions and greater primary care activity for some conditions, but not with clear differences in outpatient attendance, unplanned admission, or mortality. These observational findings do not establish that A&G caused diagnostic delay as exposure timing, case selection, coding, and unmeasured clinical complexity may explain part of the associations. Nevertheless, they support urgent evaluation of how A&G is implemented and monitored, with assessment of whole patient pathways and safety outcomes rather than referral avoidance alone.

## Copyright/License for Publication

The Corresponding Author has the right to grant on behalf of all authors and does grant on behalf of all authors, a worldwide licence to the Publishers and its licensees in perpetuity, in all forms, formats and media (whether known now or created in the future), to

i. publish, reproduce, distribute, display and store the Contribution,
ii. translate the Contribution into other languages, create adaptations, reprints, include within collections and create summaries, extracts and/or, abstracts of the Contribution,
iii. create any other derivative work(s) based on the Contribution,
iv. to exploit all subsidiary rights in the Contribution,
v. the inclusion of electronic links from the Contribution to third party material where-ever it may be located; and,
vi. licence any third party to do any or all of the above.

## Competing Interests

All authors have completed the ICMJE uniform disclosure form at http://www.icmje.org/disclosure-of-interest/ and declare: no support from any organisation for the submitted work; no financial relationships with any organisations that might have an interest in the submitted work in the previous three years; no other relationships or activities that could appear to have influenced the submitted work.

## Availability of Data and Materials

Data may be obtained from a third party and are not publicly available. The data were obtained from the Clinical Practice Research Datalink. Clinical Practice Research Datalink data governance does not allow us to distribute patient data to other parties. Researchers may apply for data access at http://www.CPRD.com/.

Code lists are openly available at https://doi.org/10.21252/8rgt-5w70

## Ethics Approval

CPRD has Health Research Authority approval to support research using anonymised patient data. This study was approved by the CPRD Research Data Governance Process (ref 25_005032; protocol made available to this manuscript’s reviewers). Under CPRD’s ethical approval from the UK Health Research Authority to support research using anonymised patient data, individual patient consent is not required as patients contributing data to CPRD cannot be identified from the data made available to researchers.

Data from CPRD adheres to the ethical principles outlined in the Declaration of Helsinki.

## Transparency

The lead author (KJM) affirms that the manuscript is an honest, accurate, and transparent account of the study being reported; that no important aspects of the study have been omitted; and that any discrepancies from the study as originally planned (and, if relevant, registered) have been explained.

## Funding

This project is funded by NIHR Health and Social Care Delivery Research (HSDR) Programme (reference number NIHR158681), and NIHR Applied Research Collaboration (ARC) West Midlands (reference number NIHR200165). CB is funded by a National Institute for Health and Care Research (NIHR) Academic Clinical Lectureship CL-2020-10-002. CJ, KPJ and CDM are part funded by the NIHR ARC West Midlands (NIHR200165). CDM is a NIHR Senior Investigator and Director of the NIHR School for Primary Care Research. CJ is an NIHR Senior Investigator. The views expressed are those of the authors and not necessarily those of the NIHR or the Department of Health and Social Care.

## Authors’ Contributions

All authors were involved in the writing - original draft and writing – review & editing of the manuscript. Attributed author roles are summarised below.

*Conceptualisation and funding acquisition:* KJM, KPJ, JB, RB, AFN, JH, RH, SLH, TH, CJ, CDM, VKW, CB

*Data curation, formal analysis and validation:* KJM, JB

*Methodology and project administration:* KJM, JB

*Supervision*: KPJ, VKW, CB

*Visualisation*: KJM

## Acknowledgments

CPRD: This study is based in part on data from the Clinical Practice Research Datalink obtained under licence from the UK Medicines and Healthcare products Regulatory Agency. The data is provided by patients and collected by the NHS as part of their care and support. The interpretation and conclusions contained in this study are those of the authors alone.

HES/ONS Data: Copyright © (2025), re-used with the permission of The Health & Social Care Information Centre. All rights reserved.

ONS Data: The interpretation and conclusions contained in this study are those of the authors alone.

PPI/ Patient Advisory Group: The authors would especially like to thank the members of the Research User Group members involved BADGER, particularly John Haines as public co-applicant on the grant, for their valuable contributions to this study. We are also thankful to Keele PPI team for their support of the public contributors.

The authors would also like to thank the BADGER Stakeholder Group, NIHR ARC West Midlands and the Study Oversight Committee for their support.

## Consent for Publication

Not applicable.

---

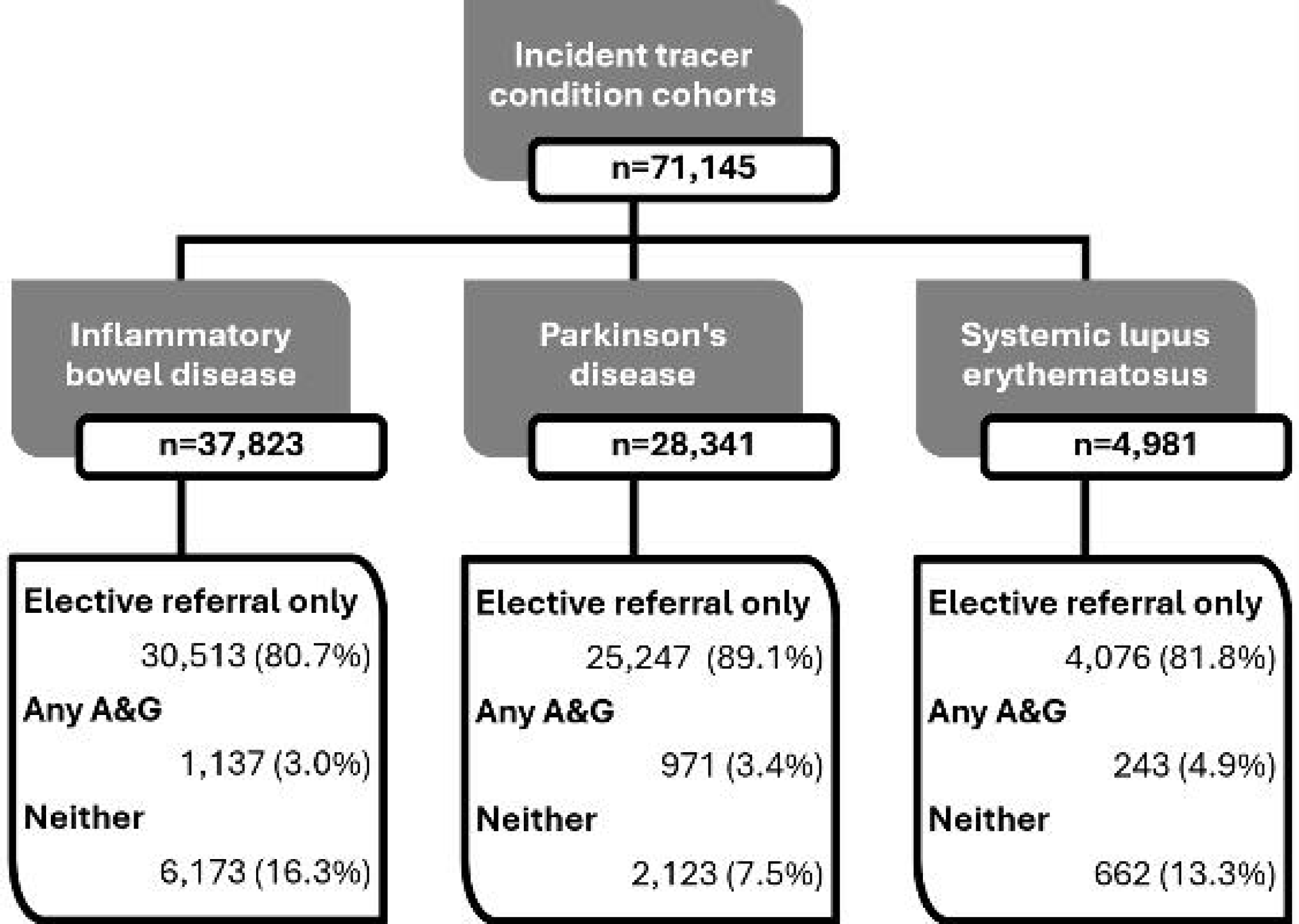

## Notes

### Competing Interest Statement

The authors have declared no competing interest.

### Author Declarations

The Clinical Practice Research Datalink (CPRD) has Health Research Authority approval to support research using anonymised patient data. This study was approved by the CPRD Research Data Governance Process (ref 25_005032; protocol made available to this manuscript′s reviewers). Under CPRD′s ethical approval from the UK Health Research Authority to support research using anonymised patient data, individual patient consent is not required as patients contributing data to CPRD cannot be identified from the data made available to researchers. Data from CPRD adheres to the ethical principles outlined in the Declaration of Helsinki.

