## Supplementary Materials for "Evaluating the impact of Advice and Guidance on diagnostic pathways and healthcare use in England: retrospective cohort study, 2015-2022"


| | |
|---|---|
| Supplementary Figure S1 | Propensity Score Balance Check: Before versus After Weighting |
| Supplementary Table S1 | Consensus Exercise for Tracer Conditions |
| Supplementary Table S2 | Definitive medical treatments for IBD, Parkinson's disease and SLE |
| Supplementary Table S3 | CODE-EHR Framework: Best practice checklist to report on the use of structured electronic healthcare records in clinical research |
| Supplementary Table S4 | Patient Characteristics including individuals without a recorded referral |

Supplementary Figure S1 Propensity Score Balance Check: Before versus After Weighting

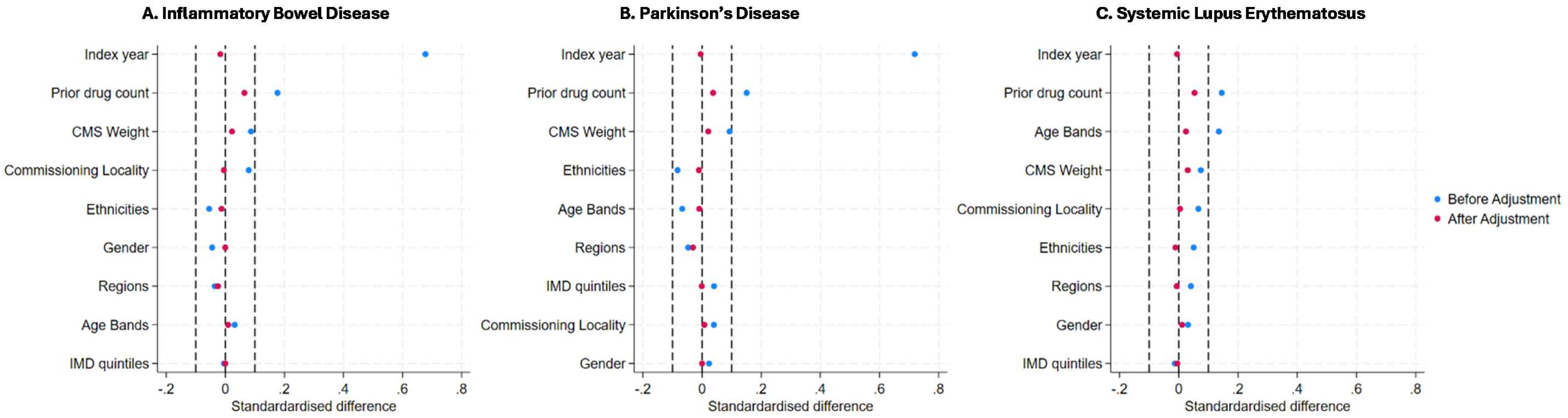


Abbreviations: CMS = Cambridge Multimorbidity Score; IMD = Index of Multiple Deprivation.

### Supplementary Table S1 Consensus Exercise for Tracer Conditions

Candidate tracer conditions were determined from patient and public involvement and engagement (PPIE), stakeholder and Study Management Group discussions, primary care clinician interviews undertaken in the funded project, relevant policy and clinical guidance, and study-team expertise. A structured, multisource consensus assessment was completed rather than a formal Delphi exercise.

Conditions were assessed against six predefined criteria: (i) frequency (operationalised using indicative prevalence of ≥0.1%); (ii) a clearly defined diagnosis coded in electronic health records; (iii) diagnosis made or confirmed in specialist care; (iv) requirement for specialist clinical management or treatment; (v) importance of timely diagnosis for patient outcomes; and (iv) representative of cross section of patient demographics.

Inflammatory bowel disease, Parkinson's disease and systemic lupus erythematosus were subsequently included as tracer conditions in the study.

| Candidate condition | Source and rationale for consideration | Assessment against criteria and feasibility | Score* | Consensus decision |
|---|---|---|---|---|
| **Inflammatory bowel disease** *(Crohn's disease; ulcerative colitis)* | Clinician interview example of A&G for patient reassurance; aligns with departmental expertise. | Frequency approximately 725 per 100,000[a] Clearly coded; specialist diagnosis and treatment; timely diagnosis important; includes children and adults; potential unplanned admissions; measurable pathway. | **6/6** | **SELECTED** **Met all criteria and provides broad age coverage of the gastroenterology referral pathway** |
| **Parkinson's disease** | Movement disorders proposed during PPIE; suspected Parkinson's disease described in a clinician interview; aligned with departmental expertise. | Frequency approximately 286 per 100,000 person-years[b] Clearly coded; specialist diagnosis and treatment; benefits from timely diagnosis; represents adults, predominantly aged 65 years and older. | **6/6** | **SELECTED** **Met all criteria and provides an older adult neurological referral pathway** |
| **Systemic lupus erythematosus** | Proposed by the Patient Advisory Group early in grant development; included in the initial CPRD feasibility exercise. | Frequency approximately 97 per 100,000[c] Clearly coded multisystem condition; specialist diagnosis and management across rheumatology, dermatology, nephrology and haematology. Lower numbers expected; treatment timing is presentation-dependent; fast-track processes may operate. | **5.5/6** | **SELECTED** **Clinically important multisystem tracer spanning several specialties; provides more diverse, middle-aged female population** |
| **Rheumatoid arthritis** | Proposed by the Patient Advisory Group; stakeholders suggested time to DMARD; clinician interview described A&G in shared care. | Met all criteria and was sufficiently frequent (approximately 779 per 100,000)[d], but an established fast-track referral pathway exists. A&G for positive rheumatoid factor or abnormal blood results may not represent confirmed disease. | **6/6** | *Not taken forward* Fast-track referral reduced suitability for comparison with routine elective referral. |
| **Spondyloarthritis** | Proposed by the Patient Advisory Group and Study Management Group. | Common (113 per 100,000)[d] and met all criteria; timely diagnosis and DMARD initiation important. Subtle presentation, but patients may pass through musculoskeletal services before rheumatology, complicating identification of the initial pathway and diagnostic interval. | **6/6** | *Not taken forward* More complex pathway to define consistently in routine data. |
| **Epilepsy** | Identified in clinician interview involving suspected epilepsy before outpatient review. | Met all criteria; sufficiently frequent (937 per 100,000)[e] and represents all ages. Established urgent or fast-track pathways, including first-seizure clinics, already exist. | **6/6** | *Not taken forward* Established fast-track pathways reduced suitability. |

| | | | | |
|---|---|---|---|---|
| **Multiple sclerosis** | Proposed by the Patient Advisory Group; policy evidence supported benefits of early diagnosis. | Met all criteria, common (190 per 100,000)[f] and represents a range of adult ages. Disease-modifying treatments are commonly hospital-issued and may be incompletely recorded in primary care, limiting measurement of definitive treatment. | **6/6** | *Not taken forward* Potentially incomplete hospital medication capture threatened the treatment outcome. |
| **Bronchiectasis** | A lung condition was suggested by a Patient Advisory Group contributor; bronchiectasis requires specialist investigation and management. | Clearly defined, sufficiently frequent (485/566 per 100,000 for males/females)[g] and likely to require specialist diagnosis, but no consistent specialist-prescribed treatment endpoint in secondary care. | **5/6** | *Not taken forward* Less clearly defined specialist-treatment endpoint. |
| **Non-malignant skin lesions** | Raised in clinician interviews; lesions were commonly associated with A&G, including image-based advice. | Infrequent (15 per 100,000)[h] although potentially important. However, heterogeneous and non-specific. Suspected melanoma and squamous cell carcinoma should use urgent suspected-cancer pathways; concerns also existed about dermatoscopy-image risk. | **5.5/6** | *Not taken forward* Definition was insufficiently specific and urgent cancer pathways complicated interpretation. |
| **Endometriosis** | Proposed by the Patient Advisory Group; supported by policy concern about diagnostic delay and gynaecology pressures. | Common (10% [190 million] of reproductive age women and girls globally)[i] and time-sensitive, but may be diagnosed or recorded following primary care investigation; definitive diagnosis may require laparoscopy. Treatment is heterogeneous and the condition represents women and girls only. | **4.5/6** | *Not taken forward* Uncertain diagnostic setting and heterogeneous treatment reduced feasibility. |
| **Motor neurone disease** | Proposed by the Patient Advisory Group following increased public awareness. | Clinically important and specialist diagnosed, but very infrequent (4.5 per 100,000 globally)[j]. Most treatment is symptom-directed and cannot be reliably attributed to motor neurone disease; riluzole is not universally prescribed. | **4.5/6** | *Not taken forward* Low numbers and no consistent definitive-treatment endpoint. |
| **Addison's disease** | Suggested by the Study Management Group as an endocrine alternative to hyperthyroidism requiring secondary-care diagnosis. | Clinically important, clearly coded and specialist managed, but uncommon (approximately 101 per 100,000; around 1-2 cases per practice)[k]. Representativeness uncertain. | **4.5/6** | *Not taken forward* Low anticipated numbers limited power and feasibility. |
| **Hyperthyroidism** | Frequently described in clinician interviews as a condition for which A&G was used. | Common (per 100,000)[l] and clearly coded, but diagnosis often occurs in primary care after thyroid-function testing. Carbimazole may also be started by GPs, independently or after specialist contact. | **3.5/6** | *Not taken forward* Primary care diagnosis and treatment conflicted with the specialist-diagnosis model. |
| **Chronic obstructive pulmonary disease / asthma** | Stakeholders identified these as common subjects of treatment-guidance requests. | Common (2,574 COPD / 5,936 asthma per 100,000)[m,n] and coded, but usually diagnosed and managed in primary care. Severe asthma treatment would yield low numbers and hospital biologics may be incompletely captured; COPD lacks a clear specialist-treatment endpoint. | **3.5/6** | *Not taken forward* Did not meet intended specialist-diagnosis and treatment criteria. |
| **Diabetes mellitus** | Diabetes management raised in a clinician interview. | Common (479 T1DM / 590 T2DM per 100,000)[o], clearly coded and representative of all ages, but diagnosis and most treatment occur in primary care. Would assess management escalation rather than specialist diagnosis; type 1 and type 2 pathways differ. | **3/6** | *Not taken forward* Did not meet specialist-diagnosis or specialist-treatment requirements. |

Abbreviations: Advice and Guidance (A&G); Clinical Practice Research Datalink (CPRD); disease-modifying antirheumatic drug (DMARD); chronic obstructive pulmonary disease (COPD); Type 1 diabetes mellitus (T1DM); type 2 diabetes mellitus (T2DM); patient and public involvement and engagement (PPIE). *Note: Scores summarise fulfilment of the six predefined criteria.

**References – Supplementary Table S1**


a. Pasvol et al. (2020) BMJ Open. 10(7):e036584
b. Parkinson's UK (2018)
c. Rees et al. (2016) Ann Rheum Dis;75:136-141
d. Scott et al. (2022) Lancet Reg Health Eur;23:100519
e. Wigglesworth et al. (2023) Seizure;105:37-42
f. Public Health England (PHE) (2020) Multiple sclerosis: prevalence, incidence and smoking status - data briefing
g. Quint et al. (2016) Eur Respir J;47(1):186-93
h. patient.info/bowens-disease
i. WHO (2023) Endometriosis. World Health Organization
j. GBD 2016 MND Collaborators. (2018) Lancet Neurol;17(12):1083-1097
k. Wass et al. (2013) Addison's Disease Self-Help Group
l. Garmendia Madariaga et al. (2014) J Clin Endocrinol Metab;99(3):923-31
m. Rayner et al. (2017) Respir Med;132:217-225
n. Mukherjee et al. (2016). BMC Med;14(1):113
o. National Diabetes Audit 2021-22, Report 1: Care Processes and Treatment Targets, Detailed Analysis Report (2013)

Supplementary Table S2 Definitive medical treatment for IBD, Parkinson's disease and SLE

| Inflammatory Bowel Disease | Parkinson's Disease | Systemic Lupus Erythematosus |
|---|---|---|
| **Steroids** | **Dopamine precursors** | **Steroids** |
| Beclomethasone | Levodopa (*benserazide/levodopa*; *carbidopa/entacapone/levodopa*; *carbidopa/levodopa*) | Methylprednisolone |
| Budesonide | Foscarbidopa | Prednisolone |
| Hydrocortisone | Foslevodopa | |
| Methylprednisolone | | |
| Prednisolone | | |
| **Anti-inflammatories** | **NMDA antagonists** | **Human Immunoglobulin (IgG)** |
| Mesalazine | Amantadine | |
| Sulfasalazine | | |
| **Immunomodulators** | **MAOB Inhibitors** | **Immunomodulators** |
| Azathioprine | Rasagiline | Azathioprine |
| Ciclosporin | Selegiline | Ciclosporin |
| Mercaptopurine | Safinamide | Cyclophosphamide |
| Methotrexate | | Hydroxychloroquine |
| | | Leflunomide |
| | | Methotrexate |
| | | Mycophenolate mofetil |
| | | Tacrolimus |
| | | Voclosporin |
| **Biologics** | **Dopamine agonists** | **Biologics** |
| Adalimumab | Bromocriptine | Belimumab |
| Golimumab | Cabergoline | Rituximab |
| Guselkumab | Pergolide | |
| Infliximab | Ramipexole | |
| Mirikizumab | Ropinirole | |
| Risankizumab | Rotigotine | |
| Ustekinumab | Apomorphine | |
| Vedolizumab | | |
| **Janus Kinase Inhibitors** | **COMT inhibitors** | |
| Filgotinib | Entacapone | |
| Tofacitinib | Opicapone | |
| Upadacitinib | Tolcapone | |
| **Other** | | |
| Etrasimod | | |
| Ozanimod | | |

Abbreviations: NMDA = N-methyl-D-aspartate; MAO-B = monoamine oxidase B; COMT = catechol-O-methyltransferase.

Supplementary Table S3 CODE-EHR Framework: Best practice checklist to report on the use of structured electronic healthcare records clinical research

Date of completion: *07/Sep/26*
Study name: *Evaluating the impact of Advice and Guidance on diagnostic pathways and healthcare use in England: retrospective cohort study, 2015-2022*

| **Item** | **Objective** | **Framework standards** | **Minimum information to provide** | **Lead Author acknowledgement** |
|---|---|---|---|---|
| 1. Dataset construction and linkage | To provide an understanding of how the structured healthcare data were identified and used. | Minimum: Flow diagram of datasets used in the study, and description of the processes and directionality of any linkage performed, published within the research report or supplementary documents.<br>Preferred: Provided within a pre-published protocol or open-access document. | (a) State the source of any datasets used.<br>(b) Comment on how the observed and any missing data were identified and addressed, and the proportion observed for each variable.<br>(c) Provide data on completeness of follow-up.<br>(d) For linked datasets, specify how linkage was performed and the quality of linkage methods. | Select one option:<br>Minimum standard not met ☐<br>Minimum standard met ☒<br>Preferred standard met ☐ |
| 2. Data fit for purpose | To ensure transparency with the approach taken, with respect to coding of the structured healthcare data. | Minimum: Clear unambiguous statements on the process of coding in the methods section of the research report.<br>Preferred: Provided within a pre-published protocol or open-access document. | (a) Confirm origin, clinical processes, and the purpose of data.<br>(b) Specify coding systems, clinical terminologies or classification used and their versions, and any manipulation of the coded data.<br>(c) Provide detail on quality assessment for data capture.<br>(d) Outline potential sources of bias. | Select one option:<br>Minimum standard not met ☐<br>Minimum standard met ☒<br>Preferred standard met ☐ |
| 3. Disease & outcome definitions | To fully detail how conditions AND outcome events were defined, allowing other researchers to identify errors and repeat the process in other datasets. | Minimum: State what codes were used to define diseases, treatments, conditions and outcomes *prior to statistical analysis*, including those relating to patient identification, therapy, procedures, comorbidities, and components of any composite endpoints.<br>Preferred: Provided within a pre-published protocol or open-access document *prior to statistical analysis.* | (a) Detailed lists of codes used for each aspect of the study.<br>(b) Date of publication and access details for the coding manual (please add to box below).<br>(c) Provide definitions, implementation logic and validation of any phenotyping algorithms used.<br>(d) Specify any processes used to validate the coding scheme or reference to prior work. | Select one option:<br>Minimum standard not met ☐<br>Minimum standard met ☒<br>Preferred standard met ☐ |
| 4. Analysis | To fully detail how outcome events were analysed and allow independent assessment of the authenticity of study findings. | Minimum: Describe the process used to analyse study outcomes, including statistical methods and use of any machine learning or algorithmic approaches.<br>Preferred: Provide a statistical analysis plan as a supplementary file, locked *prior to analyses commencing.* | (a) Provide details on all statistical methods used.<br>(b) Provide links to any machine code or algorithms used in the analysis, preferably as open-source.<br>(c) Specify the processes of testing assumptions, assessing model fit and any internal validation.<br>(d) Specify how generalisability of results was assessed, the replication of findings in other datasets, or any external validation. | Select one option:<br>Minimum standard not met ☐<br>Minimum standard met ☒<br>Preferred standard met ☐ |
| 5. Ethics & governance | To provide patients, who may or may not have given consent, and regulatory authorities the ability to interrogate the security and provenance of the data. | Minimum: Clear unambiguous statements on how the principles of Good Clinical Practice and Data Protection will be/were met, provided in the methods section of the research report.<br>Preferred: Provided within a pre-published protocol or open-access document with evidence of patient and public engagement. | (a) State how informed consent was acquired, or governance if no patient consent.<br>(b) Specify how data privacy was protected in the collection and storage of data.<br>(c) Detail what steps were taken for patient and public involvement in the research study.<br>(d) Provide information on where anonymised source data or code can be obtained for verification and further research. | Select one option:<br>Minimum standard not met ☐<br>Minimum standard met ☐<br>Preferred standard met ☒ |

| | |
|---|---|
| 6. Coding manual | DOI of publication or website address: *https://doi.org/10.21252/8rgt-5w70*<br>Date published: *01/Oct/26* |
| 7. Comments | *The approved CPRD protocol cannot be made publicly available, but was made available to reviewers of this manuscript as per CPRD guidelines.* |
| **8. Summary declaration** | One or more minimum standards not met ☐ OR **All minimum standards met** ☒<br>Number of preferred standards met: *1* / 5 |

Directions for use:

**Research team:** To complete the checklist, authors will need to consider these points during the design of the research to ensure that coding protocols and coding manuals are pre-published. Where applicable, it is advisable that all five minimum standards are met for an individual research study, whether observational or a controlled trial. If any component is not applicable to the study, the corresponding author can indicate why this is the case in the comment box.

This checklist can accompany the article as a supplementary file on submission to the journal, with the ability for readers to review responses. A comment on the meeting of standards in the text of the method section is suggested, for example; *"this study meets all five of the CODE-EHR minimum framework standards for the use of structured healthcare data in clinical research, with two out of five standards meeting preferred criteria <add reference to this CODE-EHR paper; https://doi.org/10.1136/bmj-2021-069048>"; OR "this study meets four out of five of the CODE-EHR minimum framework standards for the use of structured healthcare data in clinical research; one of the five minimum standards was not met as coding schemes were not specified prior to analysis <add reference to this CODE-EHR paper; https://doi.org/10.1136/bmj-2021-069048>."* Note, easy to complete form versions of this checklist are available in the article appendices (word and pdf versions) and at https://www.escardio.org/bigdata.

**Research appraisers (patients, clinicians, regulators, guideline task forces):** Where applicable, it is advisable that all five minimum standards are met for the research study to be considered robust.

**FURTHER DETAILS ON THE CODE-EHR FRAMEWORK:** please refer to Kotecha D, Asselbergs FW, et al; on behalf of the Innovative Medicines Initiative BigData@Heart Consortium, European Society of Cardiology, CODE-EHR international consensus group. CODE-EHR best practice framework for the use of structured electronic healthcare records in clinical research. BMJ 2022;378:e069048. doi:10.1136/bmj-2021-069048. Also published in Lancet Digit Health and Eur Heart J.

Supplementary Table S4 Patient Characteristics including individuals without a recorded referral

| Characteristic | | Inflammatory Bowel Disease (n=37,823) | | | Parkinson's Disease (n=28,247) | | | Systemic Lupus Erythematosus (n=4,981) | | |
|---|---|---|---|---|---|---|---|---|---|---|
| | | No A&G or referral recorded | Any A&G | Elective referral only | No A&G or referral recorded | Any A&G | Elective referral only | No A&G or referral recorded | Any A&G | Elective referral only |
| **Total, n** | | 6,173 (16.3) | 1,137 (3.0) | 30,513 (80.7) | 2,123 (7.5) | 971 (3.4) | 25,247 (89.1) | 662 (13.3) | 243 (4.9) | 4,076 (81.8) |
| **Female gender, n (%)** | | 2,652 (43.0) | 624 (54.9) | 15,556 (51.0) | 782 (36.8) | 347 (35.7) | 9,538 (37.8) | 532 (80.4) | 201 (82.7) | 3,384 (83.0) |
| **Median age, years (p25-p75)** | | 41 (26, 57) | 45 (28, 63) | 43 (29, 59) | 76 (69, 82) | 74 (68, 81) | 75 (69, 81) | 46 (32, 59) | 55 (41, 69) | 50 (38, 64) |
| **Age bands, n (%)** | 0-14 years | 402 (6.5) | 64 (5.6) | 1,302 (4.3) | a | a | a | a | a | 60 (1.5) |
| | 15-24 years | 959 (15.5) | 152 (13.4) | 4,127 (13.5) | a | a | a | 75 (11.3) | 08 (3.3) | 223 (5.5) |
| | 25-34 years | 1,102 (17.9) | 191 (16.8) | 5,240 (17.2) | a | a | 14 (0.1) | 89 (13.4) | 32 (13.2) | 491 (12.0) |
| | 35-44 years | 941 (15.2) | 154 (13.5) | 5,138 (16.8) | 16 (0.8) | 7 (0.7) | 138 (0.5) | 124 (18.7) | 32 (13.2) | 775 (19.0) |
| | 45-54 years | 955 (15.5) | 166 (14.6) | 4,931 (16.2) | 76 (3.6) | 37 (3.8) | 778 (3.1) | 134 (20.2) | 47 (19.3) | 843 (20.7) |
| | 55-64 years | 835 (13.5) | 148 (13.0) | 4,206 (13.8) | 228 (10.7) | 140 (14.4) | 2,861 (11.3) | 98 (14.8) | 46 (18.9) | 693 (17.0) |
| | 65-74 years | 638 (10.3) | 141 (12.4) | 3,257 (10.7) | 600 (28.3) | 306 (31.5) | 7,765 (30.8) | 72 (10.9) | 44 (18.1) | 575 (14.1) |
| | 75-84 years | 276 (4.5) | 99 (8.7) | 1,857 (6.1) | 807 (38.0) | 343 (35.3) | 10,210 (40.4) | 37 (5.6) | 27 (11.1) | 343 (8.4) |
| | 85+ years | 65 (1.1) | 22 (1.9) | 455 (1.5) | 397 (18.7) | 137 (14.1) | 3,477 (13.8) | a | a | 73 (1.8) |
| **Deprivation quintile, n (%)** | 1 - least | 960 (15.6) | 171 (15.0) | 5,283 (17.3) | 370 (17.4) | 175 (18.0) | 5,739 (22.7) | 81 (12.2) | 33 (13.6) | 627 (15.4) |
| | 2 - second least | 1,007 (16.3) | 213 (18.7) | 4,907 (16.1) | 307 (14.5) | 204 (21.0) | 4,795 (19.0) | 88 (13.3) | 45 (18.5) | 617 (15.1) |
| | 3 - mid | 1,034 (16.8) | 185 (16.3) | 4,646 (15.2) | 305 (14.4) | 163 (16.8) | 3,943 (15.6) | 108 (16.3) | 42 (17.3) | 685 (16.8) |
| | 4 - second most | 1,004 (16.3) | 190 (16.7) | 4,872 (16.0) | 272 (12.8) | 147 (15.1) | 3,372 (13.4) | 133 (20.1) | 35 (14.4) | 686 (16.8) |
| | 5 - most | 932 (15.1) | 142 (12.5) | 4,476 (14.7) | 219 (10.3) | 80 (8.2) | 2,521 (10.0) | 133 (20.1) | 46 (18.9) | 681 (16.7) |
| | Missing | 1,236 (20.0) | 236 (20.8) | 6,329 (20.7) | 658 (31.0) | 202 (20.8) | 4,877 (19.3) | 123 (18.6) | 42 (17.3) | 780 (19.1) |
| **Ethnicity, n (%)** | Asian | 508 (8.2) | 104 (9.1) | 2,270 (7.4) | 62 (2.9) | 52 (5.4) | 934 (3.7) | 111 (16.8) | 21 (8.6) | 507 (12.4) |
| | Black | 170 (2.8) | 21 (1.8) | 577 (1.9) | 41 (1.9) | 21 (2.2) | 372 (1.5) | 69 (10.4) | 25 (10.3) | 337 (8.3) |
| | Mixed/multiple | 79 (1.3) | 18 (1.6) | 365 (1.2) | a | a | 99 (0.4) | 22 (3.3) | 7 (2.9) | 68 (1.7) |
| | White | 5,138 (83.2) | 922 (81.1) | 25,071 (82.2) | 1,932 (91.0) | 852 (87.7) | 22,370 (88.6) | 444 (67.1) | 171 (70.4) | 2,909 (71.4) |
| | Other | 27 (0.4) | 22 (1.9) | 534 (1.8) | a | a | 228 (0.9) | a | 7 (2.9) | 100 (2.5) |
| | Unknown | 251 (4.1) | 50 (4.4) | 1,696 (5.6) | 85 (4.0) | a | 1,244 (4.9) | a | 12 (4.9) | 155 (3.8) |
| **Geographical** region, n (%) | London | 1,014 (16.4) | 248 (21.8) | 4,955 (16.2) | 264 (12.4) | 151 (15.6) | 3,142 (12.4) | 161 (24.3) | 56 (23.1) | 878 (21.5) |
| | Midlands/East England | 1,292 (20.9) | 262 (23.0) | 6,819 (22.3) | 600 (28.3) | 254 (26.2) | 6,104 (24.2) | 152 (23.0) | 52 (21.4) | 923 (22.6) |
| | North England | 1,766 (28.6) | 192 (16.9) | 8,403 (27.5) | 518 (24.4) | 200 (20.6) | 6,818 (27.0) | 157 (23.7) | 45 (18.5) | 1,020 (25.0) |
| | South England | 2,101 (34.0) | 435 (38.3) | 10,336 (33.9) | 741 (34.9) | 366 (37.7) | 9,183 (36.4) | 192 (29.0) | 90 (37.0) | 1,255 (30.8) |
| **Median prior consultation count, (p25-p75)** | | 16 (9, 27) | 37 (22, 58) | 27 (16, 43) | 19 (7, 33) | 45 (29, 70) | 35 (22, 53) | 20 (12, 34) | 49 (31, 71) | 37 (23, 55) |
| **Median prior medication count, (p25-p75)** | | 4 (2, 8) | 8 (4, 14) | 6 (3, 12) | 8 (4, 14) | 12 (8, 19) | 11 (6, 17) | 6 (2, 11) | 12 (7, 19) | 10 (6, 17) |
| **Any CMS Category, n (%)** | | 2,521 (40.8) | 667 (58.7) | 16,185 (53.0) | 1,968 (92.7) | 749 (77.1) | 17,826 (70.6) | 283 (42.7) | 151 (62.1) | 2,379 (58.4) |
| **Median CMS categories, (p25-p75)** | | 0 (0, 1) | 1 (0, 2) | 1 (0, 1) | 2 (1, 3) | 1 (1, 2) | 1 (0, 2) | 0 (0, 1) | 1 (0, 2) | 1 (0, 2) |
| **Median CMS Weight, (p25-p75)** | | 0.00 (0.00, 0.32) | 0.03 (0.00, 0.48) | 0.00 (0.00, 0.33) | 0.81 (0.55, 1.17) | 0.48 (0.03, 0.81) | 0.32 (0.00, 0.81) | 0.00 (0.00, 0.33) | 0.32 (0.00, 0.62) | 0.29 (0.00, 0.48) |

Abbreviations: advice and guidance (A&G); 25th-75th percentiles (p25-p75); Cambridge Multimorbidity Score (CMS); a = censored for low cell count.
